# Does the metabolic syndrome exist? The identification of cardio-metabolic profiles in a South Asian population study

**DOI:** 10.1101/19012195

**Authors:** Lisa Harber-Aschan, Ioannis Bakolis, Nicholas Glozier, Khalida Ismail, Kaushalya Jayaweera, Gayani Pannala, Carmine Pariante, Fruhling Rijsdijk, Sisira Siribaddana, Athula Sumathipala, Helena M.S. Zavos, Patricia Zunszain, Matthew Hotopf

**Affiliations:** Department of Psychological Medicine, Institute of Psychiatry, Psychology, and Neuroscience, King’s College London, UK; Department of Biostatistics and Health Informatics, Department of Health Services and Population Research, Institute of Psychiatry Psychology and Neuroscience, London, UK; Centre for Implementation Science, Health Services and Population Research Department, Institute of Psychiatry Psychology and Neuroscience, London, UK; Brain and Mind Centre, University of Sydney, Australia; Institute for Research and Development, Colombo, Sri Lanka; Stress, Psychiatry and Immunology Laboratory, Institute of Psychiatry, Psychology & Neuroscience, King’s College London, UK; Social Genetic and Developmental Research Centre, Institute of Psychiatry, Psychology & Neuroscience, King’s College London, UK; Department of Medicine, Faculty of Medicine & Allied Health Sciences, Rajarata University of Sri Lanka, Sri Lanka; Research Institute for Primary Care & Health Sciences, Faculty of Medicine & Health Sciences, Keele University, UK; Department of Psychology, Institute of Psychiatry, Psychology & Neuroscience, King’s College London, UK; South London and Maudsley NHS Foundation Trust, London, UK

## Abstract

**OBJECTIVE:** Research testing the validity of the metabolic syndrome (MetS) as a clinical construct associated with cardiovascular disease risk has produced inconsistent results. This study tested the existence of the MetS, explored alternative cardiometabolic risk characterisations, and examined the relative influence of genetic and environmental factors in a South Asian sample.

**RESEARCH DESIGN AND METHODS:** Data came from the Colombo Twin and Singleton follow-up Study, CoTaSS-2 (N=3969). Latent class analysis tested the clustering of MetS indicators (waist circumference, high-density-lipoprotein cholesterol (HDL-C), triglycerides (TG), blood pressure, fasting plasma glucose (FPG), medications and diabetes). Regression analyses tested cross-sectional associations between identified latent classes and covariates. Structural equation modelling estimated genetic and environmental influences on these classes. Analyses were stratified by gender (n=1681 men, n=2288 women).

**RESULTS:** Three classes were identified in men: 1) “Healthy” (52.3%), 2) “Central obesity, high TG, high FPG” (40.2%), and 3) “Central obesity, high TG, diabetes” (7.6%). Four classes were identified in women: 1) “Healthy” (53.2%), 2) “Very high central obesity, low HDL-C, raised FPG” (32.8%), 3) “Very high central obesity, diabetes” (7.2%) and 4) “Central obesity, hypertension, raised FPG” (6.8%). Older age in men and women, and high socioeconomic status in men, was associated with cardiometabolic risk categories, compared to the “Healthy” classes. In men, individual differences in cardiometabolic class membership were due to environmental effects. In females, genetic differences significantly predicted class membership.

**CONCLUSIONS:** The findings did not support the MetS construct. Instead, distinct clinical profiles were identified in men and women, suggesting different aetiological pathways.

---

The metabolic syndrome (MetS) is a clinical construct of risk factors for cardiovascular disease (CVD) and other cardiometabolic diseases, including central obesity, a poor lipid profile, blood pressure, and hyperglycaemia (1). Whilst MetS increases CVD risk, beyond its component parts (2, 3), the MetS components’ relative importance, measurement, and thresholds cut-offs are debated (4). More fundamental questions regarding the syndrome’s aetiology, clinical utility, and validity, have also been raised. Studies using statistical clustering methods to test the construct validity of MetS have produced inconsistent results (5). Whilst some have found support for a cohesive MetS construct (6–8), most find 3-4 clusters, and the clinical composition often varies substantially between ethnicities, countries, gender, and ages (5, 9–16). This heterogeneous evidence base speaks against the “natural” clustering of components, united by a single underlying mechanism (1), and the universality of MetS. Rather, it suggests that cardiometabolic risk profiles are specific to gender, age and culture, with potentially different aetiological pathways carrying different cardiometabolic risks.

As cardiometabolic diseases become increasingly prominent public health challenges in low- and middle-income countries, there is a need to understand the variations in cardiometabolic risk that may be different to high income settings. This is particularly true for South Asian populations where the morbidity of diabetes and CVD is steadily increasing, following a rapidly changing landscape of urbanisation and Westernisation (17). As a postulated precursor of these conditions, validating MetS in a South Asian population could identify an important risk group. Understanding the aetiology of cardiometabolic risk in South Asian populations also warrants estimating of genetic contributions, particularly in the context of the “thrifty” genotype hypothesis proposing extreme sensitivity to metabolic dysregulation in face of obesogenic challenges in these populations (18). This study used a Sri Lankan population study with a genetically sensitive design to: 1) Statistically test the MetS; 2) explore alternative characterisations of cardiometabolic risk, and 3) examine the relative influence of genetic and environmental factors contributing to cardiometabolic risk profiles. We hypothesised that a cohesive MetS latent class would emerge, with elevated cardiometabolic risk for all MetS components.

## Research Design and Methods

### Sample

A representative population sample from the Colombo district in Sri Lanka was used (the Second Colombo Twin and Singleton Study, CoTaSS-2). CoTaSS is a two-wave cohort study of mental and physical health of twins and singletons, CoTaSS −1 taking place in 2007 initially; follow-up for CoTaSS −2 taking place 2012-2015 (retention rate 76.4%). We used cross-sectional data from respondents from CoTaSS-2 (N=3476). Details of the recruitment and data collection procedures have been described elsewhere (19).

### Measures

#### Metabolic syndrome (metS)

MetS was calculated according to the International Diabetes Federation (IDF) and National Cholesterol Education Programme Adult Treatment Panel (NCEP ATP III) definitions (20, 21). The IDF criteria requires central obesity (South Asian specific thresholds of ≥90cm for men and ≥80cm in women) and at least two additional components: 1) elevated blood pressure (BP; systolic≥130 mmHg and/or diastolic≥85 mmHg, or hypertensive treatment), 2) elevated plasma triglycerides (TG; ≥1.70 mmol/L), 3) low high-density lipoprotein cholesterol (HDL-C; <1.03 mmol/L in men, <1.29 mmol/L in women), and 4) elevated FPG (≥5.6 mmol/L or diabetes treatment). The revised NCEP ATP III criteria accepts any three components, without holding central obesity as essential. The MetS indicators used in the analysis thus included: waist circumference (WC), TG, HDL-C, systolic blood pressure (SBP), and FPG, and self-reported diabetes diagnoses or medication. The non-computerised calculation of the Homeostasis Model Assessment (HOMA) score estimated insulin resistance (22, 23). Hypertension, heart conditions, hypertensive medication, and cholesterol medication were self-reported. Additional clinical measures indicative of cardiometabolic risk were obtained from blood and urine samples (19).

#### Socio-demographic measures

A questionnaire captured sex, age, marital status (married vs. not married), ethnicity (Sinhala vs. Tamil/Muslim/Other), urbanicity (Urban vs. rural/mixed/outside Colombo), education (≤ Grade 5, Grade 6-O/Ls, ≥ A/Ls), occupational class (Managers/Professionals, Skilled manual/non-manual workers, Elementary occupations, Not in employment), and perceived financial strain (Low: living comfortably/doing alright, Moderate: just about getting by, High: difficult/very difficult to make ends meet).

#### Health behaviours

Physical activity was measured by an internationally validated screen, capturing low, moderate and high physical activity (24). A culturally adapted diet-questionnaire asked about food groups consumed (25). A continuous diet risk variable ranging from 0-5 was constructed by adding binary variables indicating under-consumption (fruit, vegetables) and over-consumption (deep fried foods, salty snacks, desserts). The Alcohol Use Disorder Identification Test (AUDIT) (26) indicated hazardous (8-point cut-off) and harmful (16-point cut-off) alcohol consumption. Self-reported tobacco smoking contrasted current smokers against non-current smokers.

### Statistical Analyses

Latent class analysis (LCA) was performed in MPlus 7, and descriptive and inferential statistics were performed in Stata 14 (27, 28). For the purposes of identifying the cardiometabolic classes, the sample was treated as a population sample, using the Stata command *svy* to account for clustering within twin-pairs, and the *type=complex* command to compute robust standard errors in MPlus. Analyses were stratified by sex. LCA addressed the aims to 1) statistically test the MetS, and (2) explore alternative characterisations of cardiometabolic risk. LCA is a data-driven approach which examines the clustering of individuals, based on their observed continuous or categorical clinical characteristics. It also considers variation in continuous clinical indicators, in contrast to MetS combination approaches using defined cut-offs. LCA therefore allows for testing the MetS, and investigating alternative cardiometabolic clusters. The indicators contributing to the LCA analysis included continuous measures of: WC, diastolic BP, systolic BP, FPG, HDL-C and TG; and binary variables of: hypertension medication, cholesterol medication, and diabetes/diabetes medication. Error terms for diastolic and systolic BP were correlated to account for violation of the independence assumption. TG and FPG measures were log-transformed. Age was entered as an auxiliary variable. Maximum-likelihood methods estimated the model parameters.

Consistent with conventional LCA practice we forced a 2-class solution, incrementally extracting additional classes. Model fit indices included: the Akaike Information Criteria (AIC) (29), Bayesian Information Criteria (BIC) (30), sample size adjusted BIC (SABIC) (31), entropy (32) (>0.80 indicates low uncertainty in classification), and the Lo–Mendell– Rubin likelihood ratio test (LMR–LRT) (33). Descriptive statistics estimated means and percentage prevalence estimates with 95% CIs to describe classes according to clinical variables contributing to the model, additional clinical measures, socio-demographic and socio-economic characteristics and health behaviours. Unadjusted and adjusted associations were explored between the latent classes and socio-demographic characteristics with multinomial regression analysis, estimating relative risk ratios (RRR) and 95% CIs. Alcohol and tobacco use were only examined in men, as these were rare among women (>1%) (19).

#### Twin analysis

Aim 3) (to examine the relative influence of genetic and environmental factors contributing to cardiometabolic risk profiles) was addressed with structural equation twin model-fitting. Dichotomous variables were generated to address the nominal nature of the class structure. A liability threshold model was fitted to each dichotomous class variable for males and females separately. This model assumes the observed proportions of e.g. variable ‘1 not 1’ to reflect an underlying normally-distributed liability that represents an individual’s propensity of belonging to that class, with the threshold indicating the prevalence of the class. A pair’s joint distribution is similarly represented by a bivariate normal distribution. Differential tetrachoric correlations in MZ and DZ twin pairs provided the power to decompose the variance of the latent liability (L) of class membership into three independent sources: additive genetic (A), shared environment (C) which makes individuals in the same family similar, and non-shared unique environment including measurement error (E) which makes individuals in the same family different (34). Model-fitting was conducted in OpenMx with raw data maximum likelihood estimation (35). Opposite-sex twin pairs could not be used since the classes were qualitatively different across males and females, however, singletons were included to inform the threshold (and the effect of age on this threshold) representing the prevalence of the classes in the population.

## Results

The mean age of men was 42 (N=1509), and 43 in women (N=1967; Table 1). Similar proportions of men and women were married (73%) and of Sinhala ethnicity (93%). Most lived in urban areas and had at least some secondary education.

**Table 1.** Descriptive table of sample and latent cardiometabolic classes.

|  | Men N=1509 |  | Women N=1967 |  |
| --- | --- | --- | --- | --- |
|  | n | %<br>/mean | n | %<br>/mean |
| **Mean age** | 1509 | 42.0 | 1967 | 43.2 |
| **Age categories** |  |  |  |  |
| 19-34 | 557 | 36.9 | 611 | 31.1 |
| 35-49 | 502 | 33.3 | 723 | 36.8 |
| 50-59 | 235 | 15.6 | 361 | 18.4 |
| 60+ | 215 | 14.2 | 272 | 13.8 |
| **Marital status** |  |  |  |  |
| Married | 1096 | 73.5 | 1417 | 72.6 |
| Not married | 395 | 26.5 | 536 | 27.4 |
| **Ethnicity** |  |  |  |  |
| Sinhala | 1391 | 93.4 | 1806 | 92.5 |
| Tamil/Muslim/Other | 99 | 6.6 | 147 | 7.5 |
| **Urbanicity** |  |  |  |  |
| Urban | 893 | 59.2 | 1183 | 60.1 |
| Rural, mixed, or outside Colombo | 616 | 40.8 | 1183 | 39.9 |
| **Education** |  |  |  |  |
| ≤ Grade 5 | 97 | 6.5 | 178 | 9.2 |
| Grade 6 - O/Ls | 947 | 63.9 | 1179 | 60.6 |
| ≥ A/Ls | 439 | 29.6 | 587 | 30.2 |
| **Occupational class** |  |  |  |  |
| Managers/Professionals | 136 | 9.2 | 130 | 6.7 |
| Skilled manual/non-manual workers | 902 | 60.9 | 481 | 24.8 |
| Elementary occupations | 193 | 13.0 | 95 | 4.9 |
| Not in employment | 250 | 16.9 | 1233 | 63.6 |
| **Financial strain** |  |  |  |  |
| Low | 1172 | 78.6 | 1418 | 72.6 |
| Moderate | 192 | 12.9 | 293 | 15.0 |
| High | 127 | 8.5 | 241 | 12.3 |
| **Latent cardiometabolic classes in men** |  |  |  |  |
| Class 1: Healthy<br>*Healthy WC, Healthy TG, Healthy HDL-C, Healthy BP Healthy FPG* | 798 | 52.3 |  |  |
| Class 2: WC, TG, FPG<br>*High WC, High TG, Healthy HDL-C, Healthy BP, Raised FPG* | 606 | 40.2 |  |  |
| Class 3: WC, TG, Diabetes<br>*High WC, High TG, Healthy HDL-C, Healthy BP, Very high levels of FPG* | 114 | 7.6 |  |  |
| **Latent cardiometabolic classes in women** |  |  |  |  |
| Class 1: Healthy, WC<br>*High WC, Healthy TG, Healthy HDL-C, Healthy BP, Healthy FPG* |  |  | 1046 | 54.2 |
| Class 2: Obese, HDL-C, Treated BP, FPG<br>*Very high WC, Borderline raised TG, Low HDL-C, Healthy BP but BP medication, Raised FPG* |  |  | 646 | 32.8 |
| Class 3: WC, Diabetes<br>*Very high WC, Borderline raised TG, Healthy HDL-C, Borderline raised BP, Very high levels of FPG* |  |  | 142 | 7.2 |
| Class 4: WC, Untreated BP, FPG<br>*High WC, Healthy TG, Healthy HDL-C, High BP but no BP medication, Raised FPG* |  |  | 133 | 6.8 |

WC, waist circumference; TG, triglyceride; HDL-C, high density lipoprotein cholesterol; BP, blood pressure; FPG, fasting plasma glucose
Note: numbers may not add up due to missing data

Guided by model fit statistics and interpretability, we opted for the 3-class solution in men, and the 4-class solution in women (see Supplemental Table S1 for fit statistics). In men, the AIC, BIC and SABIC improved with the introduction of additional classes, but the LMR-LRT *p*-value indicated that no further model fit improvements were gained beyond 3 classes. Entropy dropped from 2 to 3 classes, suggesting poorer parsimony, but remained acceptable. Despite poorer parsimony, we opted for the 3-class solution as the classes were more clinically meaningful upon inspection. In women, the AIC, BIC and SABIC improved with additional classes, and all models had good entropy (>0.8) indicating clearly delineated classes. Model fit improved until the 4-class solution.

Fig. 1 presents the latent classes by MetS components in men and women (see Supplemental Table S2 for exact means). Approximately half of the men were in a “Healthy” class (Class 1), while 40.2% were in Class 2 characterised by high WC, high TG and raised FBG, and 7.6% of men in Class 3 (7.6%) had substantially raised levels of FBG, and high WC and TG. Men in Classes 2 and 3 were similar on most MetS components, but Class 2 had FPG levels just above threshold (5.8 mmol/L), while men in Class 3 had nearly twice as high FBG levels (11.5 mmol/L).

**Figure 1a-e.**
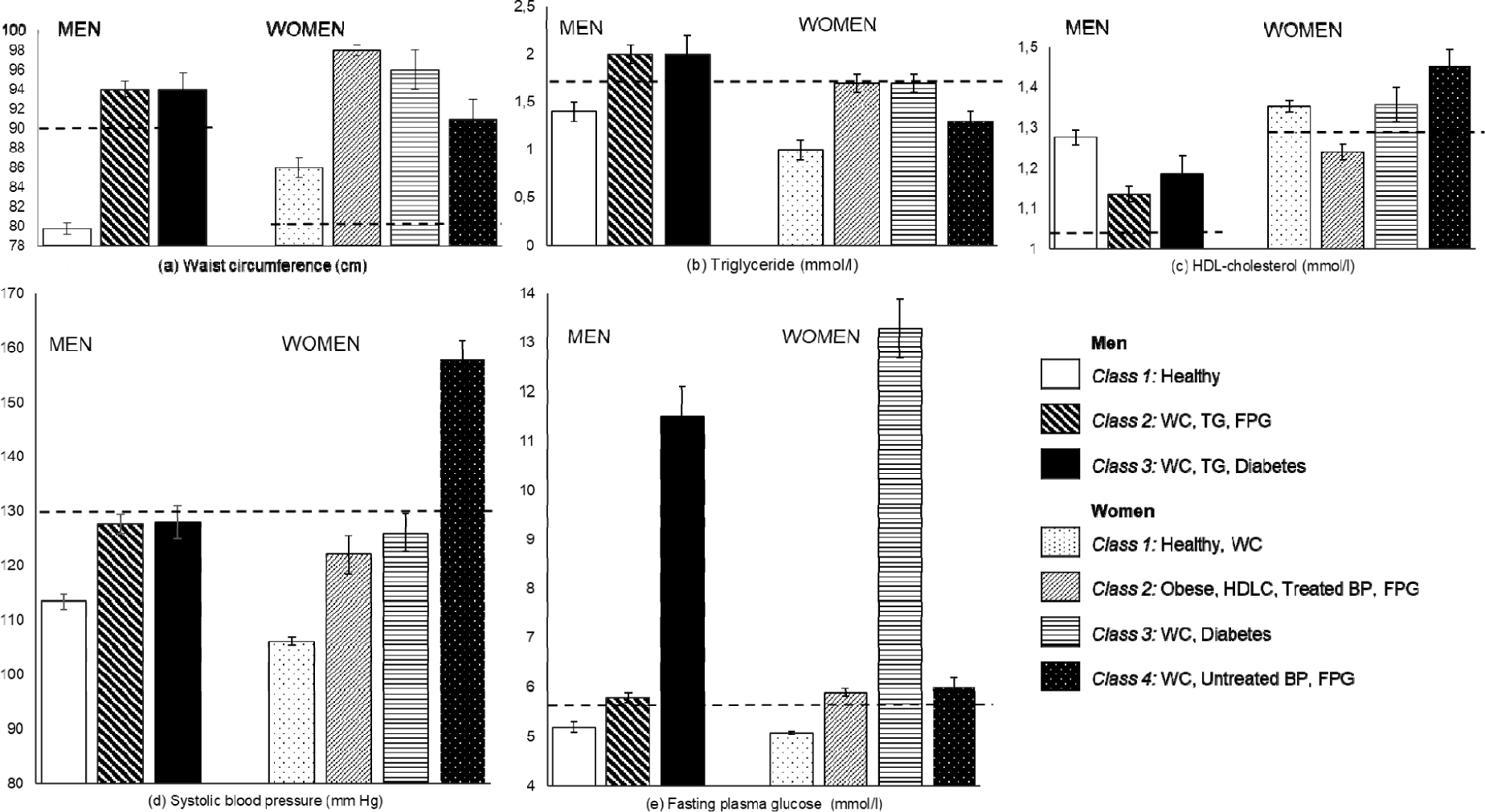
Cardiometabolic classes by metabolic syndrome components. The horizontal lines indicate risk thresholds defined by the IDF and NCEP ATP III criteria. Higher values indicate poorer cardiometabolic function with the exception of HDL-cholesterol where values below the line indicate poor cardiometabolic health. Systolic blood pressure is presented in (d); diastolic blood pressure distributions were comparable (not shown).

Fifty-four percent of women belonged to a “Healthy” class (Class 1), with WC above threshold (85.6 cm vs. 80.0 cm, Fig. 1), but an otherwise healthy cardiometabolic profile. Low HDL-C specifically affected women in Class 2 (32.8%), who also had raised TG and FPG. Whilst women in Class2 had the lowest BP levels, it seems that they have well-managed hypertension, given that a high proportion reported BP medication (Supplemental Table S2). Substantially raised FPG levels were characteristic for women in Class 3 (7.2%), while untreated hypertension was a distinct clinical characteristic of women in Class 4 (6.8% of the sample). The mean WC was above the risk threshold for all female Classes, but particularly high in Class 2.

Class 1, the “Healthy” class, and Class 3, the “Diabetes” class, were comparable in their clinical profiles and proportions between men and women. There were also notable gender differences. Women’s WC in all classes exceeded the sex-specific cut-off more than for men. TG levels were above risk threshold in male Classes 2 and 3, whilst only borderline threshold for women in Classes 2 and 3. None of the male classes were characterised by poor HDL-cholesterol profiles or high BP, while this specifically affected women in Class 2 and Class 4.

Table 2 presents adjusted associations between socio-demographic characteristics and health behaviours with the cardiometabolic classes, compared to Class 1 (see Supplemental Table S3 and Table S4 for prevalence distributions and unadjusted analyses). In men, there was strong evidence that older age and being married increased the risk of membership in Class 2 and 3, compared to Class 1. Men with no educational qualifications and in elementary occupations were at substantially lower risk of placement in Classes 2 and 3, relative to the “Healthy” Class 1 (Table 2). U-shaped trends were observed for physical activity and alcohol use in men, such that moderate - but not low - physical activity, and hazardous - but not harmful - alcohol use was associated with risk of placement in Class 2 and 3, relative to the reference. In women, there was strong evidence for age being a determinant of cardiometabolic class membership; older age being particularly strongly associated with placement in Class 4. For women, evidence suggested that education, occupational class, financial strain, physical activity and diet did not determine class membership. Adjusted regression analyses exploring differences between the cardiometabolic classes, indicated that age was the only covariate which distinguished classes from each other. In men, older age was associated with increased risk of membership in Class 3 over Class 2 (RRR=1.04 (1.02-1.06), p<0.001). Similarly in women, older age was associated with increasing risk of membership across the classes (Class 3 relative to Class 2: RRR=1.02 (1.00-1.03) p<0.035; Class 4 relative to Class 2: RRR=1.12 (1.09-1.15), p<0.001; Class 4 relative to Class 3: RRR=1.10 (1.08-1.14), p<0.001; analyses not shown).

**Table 2.**
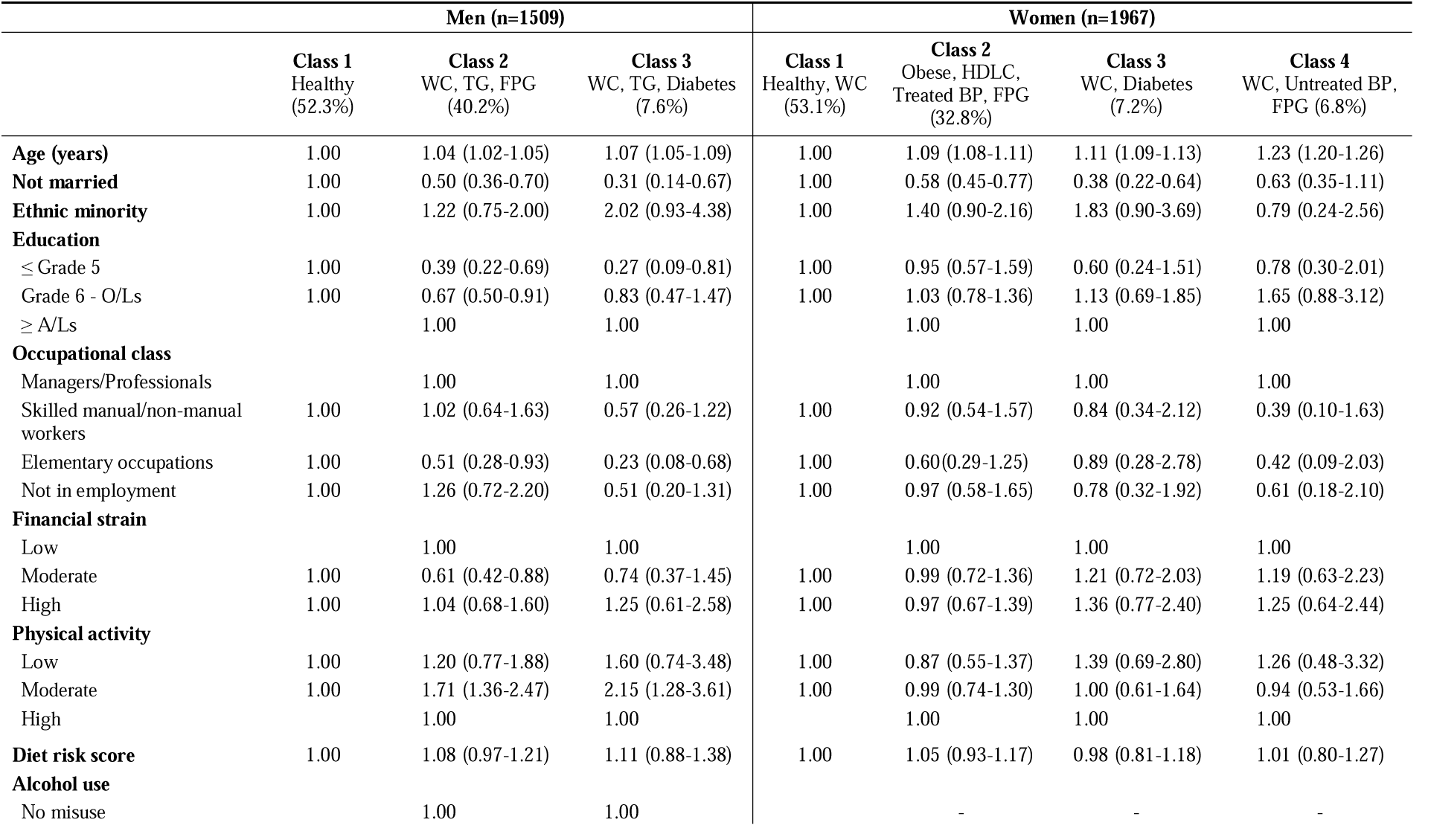
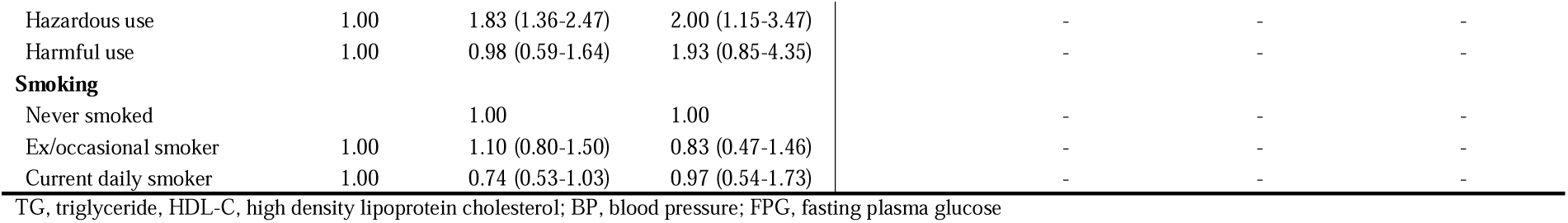
Associations between latent cardiometabolic classes and socio-demographic characteristics and health behaviorus (relative risk ratios, 95% CI, p-value; adjusted for all variables presented in the table)

Approximately half of men in Class 2 and 58.8-71.9% of men in Class 3 met the MetS criteria, while very few men in Class 1 did so (Table 3). In women, MetS was prevalent among approximately 10%, 70%, 80% and 65% of women in Classes 1-4, respectively. The distribution of other cardiometabolic risk factors indicated clinical profiles which were broadly consistent with the characterisations of the cardiometabolic classes (Supplemental Table S5). Men and women in respective Class 1 identified as “healthy” had healthy BMIs and good overall clinical profiles. FPG measures were consistent with measures of HbA1c and insulin resistance, and cholesterol measures were broadly consistent with HDL-C levels, and self-reported hypertension was highest in female Class 4 (the untreated hypertension class). Notably, microalbumin, ACR and CRP levels mirrored FPG levels and were substantially higher among in men and women in Class 3, characterised by diabetes with adverse renal function and inflammation.

**Table 3.** Overlap between latent cardiometabolic classes and metS (column %, 95% CI)

|  | Men (n=1509) |  |  | Women (n=1967) |  |  |  |
| --- | --- | --- | --- | --- | --- | --- | --- |
|  | Class 1<br>Healthy<br>(52.3%) | Class 2<br>WC, TG, FPG<br>(40.2%) | Class 3<br>WC, TG, Diabetes<br>(7.6%) | Class 1<br>Healthy, WC<br>(53.1%) | Class 2<br>Obese, HDLC,<br>Treated BP, FPG<br>(32.8%) | Class 3<br>WC, Diabetes<br>(7.2%) | Class 4<br>WC, Untreated BP,<br>FPG (6.8%) |
| MetS (IDF) (col %) | 0.6 (0.3-1.5) | 42.4 (38.5-46.4) | 58.8 (49.0-67.9) | 9.8 (8.1-11.8) | 73.4 (69.8-76.7) | 79.6 (71.8-85.7) | 65.9 (57.5-73.4) |
| MetS (ATP III) (col %) | 3.3 (2.3-4.8) | 51.1 (47.1-55.0) | 71.9 (62.6-79.7) | 9.7 (8.0-11.7) | 73.7 (70.2-77.0) | 81.0 (73.3-86.9) | 65.9 (57.4-73.5) |

MetS, metabolic syndrome

For the twin analyses, sample size did not permit decomposing class-membership variance for male Class 3 (N=114) and females Class 4 (N=142) due to few concordant pairs. For the other classes, individual differences in class membership in males was equally due to the two components that make up familial effects: genetic and shared-environmental variance (Table 4). There was, however, no power to establish the significance of these components independently, only their sum (familial effects). For females, in contrast, genetic effects determined membership of all Classes, with high heritability estimates (70-78%). The small DZ twin correlations in females suggested fitting a model with non-additive genetic effects rather than C (shared environment). However, the analysis was under-powered to estimate two different genetic sources, and we primarily aimed to estimate overall heritabilities (34, 35).

**Table 4.** Twin correlations and ACE decomposition (with 95% CI) of class membership for males and females.

|  | r<sub>MZ</sub> | r<sub>DZ</sub> | h<sup>2</sup> | c<sup>2</sup> | e<sup>2</sup> |
| --- | --- | --- | --- | --- | --- |
| Males |  |  |  |  |  |
| Class 1: Healthy | .76 (.61/.86) | .54 (.30/.73) | .40 (.00/.86) | .40 (.00/.76) | .20 (.11/.32) |
| Class 2: WC, TG, FPG | .65 (.48/.78) | .42 (.15/.64) | .46 (.00/.80) | .23 (.00/.67) | .31 (.19/.47) |
| Females |  |  |  |  |  |
| Class 1: Healthy, WC | .75 (.61/.85) | .05 (-.24/.34) | .73 (.35/.92) | .00 (.00/.20) | .28 (.17/.43) |
| Class 2: Obese, HDLC, Treated BP, FPG | .66 (.50/.79) | .13 (-.14/.37) | .70 (.39/.81) | .00 (.00/.27) | .30 (.19/.44) |
| Class 3: WC, Diabetes | .73 (.44/.90) | .14 (-.41/.62) | .78 (.08/.92) | .00 (.00/.61) | .22 (.08/.46) |

Note: h<sup>2</sup> = standardized genetic effects; c<sup>2</sup> = standardized shared-environmental effects; e<sup>2</sup> = standardized unique-environmental effects. r<sub>MZ</sub> = MZ twin correlation; r<sub>DZ</sub> = DZ twin correlation

## Conclusions

This study found limited support for a unified MetS construct in a Sri Lankan population sample. Model solutions with more than two classes fitted the data better, and none could be characterised as a distinct MetS class. Instead, we found distinct clinical profiles, specific to men and women, with high cardiometabolic risk on some components, while others within the same class were within healthy ranges or only slightly raised. Specifically, for women, the identified classes had distinct cardiometabolic risk profiles, such Class 3 characterised by very high FBP, and Class 4 characterised by very high BP. For men, the cardiometabolic risk classes were more comparable across components, but distinguished by substantially raised FPG values in Class 3. Class 2 in women had the closest match to a congruent MetS construct, with very high WC, low HDL-C, treated hypertension, and raised TG and FPG. However, substantial heterogeneity across indicators was observed: whilst WC and HDL-C were substantially over the threshold limit, the other components were not comparably high as would be expected if they were to cluster “naturally”, and were driven by a single underlying mechanism (1).

Support for different versions of MetS classes have been found in other LCA studies. For example, a two-class solution was found in men and women of Hispanic ethnicity in the US (7); a study of Japanese-American men found a distinct MetS class in a three-class solution (11); a study of an Iranian population sample observed a MetS class among four identified classes (13); and a US study identified a MetS class in an ethnically diverse population sample (14). However, the identified MetS classes did not score highly consistently across all components (13, 14), certain components were excluded for the purposes of achieving parsimony (7), and additional distinct cardiometabolic risk classes were typically found with high risk on some components, consistent with our findings (11, 13, 14). Furthermore, the MetS classes identified varied in size, and in terms of in the prominence of MetS components contributing to them, and differed by gender (14). Our results contribute to this heterogenous evidence base, indicating that there is substantial variation in the expression of MetS component clustering, and limited evidence for a single underlying MetS construct. Research directly comparing MetS component clustering between countries in Europe also observed substantial cross-cultural variation, supporting this interpretation (12).

Our findings are consistent with past studies using factor analyses of Asian population samples which also identified multiple clusters of MetS components, which were gender-specific (9, 10). Specifically, a study using a multi-ethnic Asian household survey which oversampled ethnic minorities (N=1957 men, N=2308 women) found that hypertension had a weak association with FPG and insulin resistance in Indian women (10), consistent with the distinct female hypertension classes with relatively low FPG in our study. Our study extends this evidence base of MetS clusters in Asian populations, using a latent class analysis approach which allows for using both binary and continuous variables, thus including those with diabetes and hypertension medication, rather than excluding these individuals as past studies have done (10). Our observation that the largest cardiometabolic risk class in women was characterised by low HDL-C is also consistent with a small study which found that low HDL-C was particularly prominent in Indian women living in the US (36) (N=226 men and women).

Low SES was protective of membership in Classes 2 and 3 in men, suggesting that lifestyle factors of men with higher SES in Sri Lanka is associated with greater cardiometabolic risk (37). However, this association held whilst adjusting for health behaviours including diet, alcohol consumption, exercise and smoking, suggesting that either unmeasured aspects of high SES in men puts them at increased cardiometabolic risk, or the lifestyle questions were prone to misclassification. The lack of an association between SES and cardiometabolic risk in women suggests that the potentially harmful effects of men’ high SES environments may not extend to women in Sri Lanka; the difference in lifestyles between men with low and high SES being greater, compared to women. Limited environmental variation may also explain why genetic effects were only observed in women given that it allowed greater opportunity for genetic effects, and why only age distinguished latent classes in women, potentially reflecting a progression cardiometabolic risk over women’s life course, or alternatively, cohort effects.

### Strengths & limitations

Although all respondents could self-report medications, direct assessment was incomplete for all MetS components, thus, selective medication measurement may have influenced group membership. The cross-sectional data limits the causal inferences made regarding environmental and behavioural risk factors. Nevertheless, this is the first study to test the clustering of MetS components in a Sri Lankan population sample, which provides a detailed description of local risk profiles, and also the first to assess metS component clustering in a large representative population sample with a genetically sensitive in design. These findings ought to be replicated in other datasets before drawing conclusions about the classes’ generalizability. Furthermore, we only included MetS components: future research should consider including other clinical measures to improve the cardiometabolic risk characterisation and prediction.

### Implications

We found that cardiometabolic risk components that ought to cluster together if they belonged to an underlying and unifying MetS construct, do not – a findings which has substantial support in the existing literature. This suggests that cardiometabolic risk cannot be universally captured in a construct such as MetS, but that cardiometabolic risk is culturally-, ethnicity- and sex-specific with distinct aetiological pathways. Despite the accumulating evidence of heterogeneity in cardiometabolic risk profiles, fundamental questions about the existing definition of MetS are rarely raised (5), but the discussions tends to encourage exploration of “MetS sub-types” instead (38). However, it is arguably inappropriate speak of sub-types when the evidence for a unifying MetS construct is limited.

Our results question the clinical usefulness of MetS, given that no class perfectly overlapped with MetS. Even for Class 3 which captured 80% of women with MetS, this nevertheless leaves 1 in 5 women with alarmingly high FBP levels unidentified by the MetS. Simultaneously, MetS misclassifies 1 in 10 women with a seemingly healthy cardiometabolic profile (Class 1). This suggest that MetS at best is a blunt tool for capturing cardiometabolic risk; at worst fails to identify a substantial proportion of people who could benefit from clinical intervention, whilst erroneously labelling a substantial minority of healthy women. Evidence also suggests that MetS is imprecise in terms of risk prediction, as only a handful of those with MetS develop CVD or diabetes. Other risk prediction tools or, indeed, specific clusters or individual MetS components may therefore predict CVD and cardiovascular ageing better (39, 40).

Whilst cross-sectional data does not permit determining whether the identified classes are superior for capturing cardiometabolic risk over the MetS, the results nevertheless bear some immediate clinical implications. The risk profiles could potentially be more meaningful to clinicians, as simple demographic indicators such as age and could inform which cardiometabolic risk factors to prioritise in terms of screening and targeting for intervention. For public health purposes, the classes indicate the greatest local population needs. Furthermore, the observation that the WC of women in the “healthy” class was substantially over the advised cut-off suggests that this recommendation for female central obesity may be substantially lower than necessary.

### Conclusion

In this Sri Lankan population sample distinct clinical profiles emerged suggesting that cardiometabolic risk is more complex than a single underlying MetS construct.

## Data Availability

The data used for the purposes of this study are available upon reasonable request and require the approval of the COTASS steering committee.

## Author Contributions

M.H. and A.S. were the joint principal investigators, conceptualised the study design and data collection and gained funding. L.H.A. generated the study aims and hypothesis, performed the latent class analyses, descriptive analyses and regression analyses, and drafted the article. I.B. provided support for the latent class analysis, and interpretation of results. F.R. and H.M.S.Z. are responsible for the twin analysis and their interpretation; L.H.A. takes responsibility for the integrity of the data and the accuracy of all other data analyses. All authors critically revised the article, and approved the final version.

## Prior Presentation

This work was presented as a conference abstract at the 22nd World Congress of the World Association for Social Psychiatry, New Delhi, India November 30^th^ – December 4^th^ 2016.

